# Is Point-of-Care testing feasible and safe in care homes in England? An exploratory usability and accuracy evaluation of a point-of-care polymerase chain reaction test for SARS-COV-2

**DOI:** 10.1101/2020.11.30.20240010

**Authors:** Massimo Micocci, Adam L Gordon, Mikyung Kelly Seo, A. Joy Allen, Kerrie Davies, Dan Lasserson, Carl Thompson, Karen Spilsbury, Cyd Akrill, Ros Heath, Anita Astle, Claire Sharpe, Rafael Perera, Gail Hayward, Peter Buckle, on behalf of the CONDOR Study Group

**Affiliations:** NIHR London In Vitro Diagnostics Co-operative, Dept of Surgery and Cancer, Faculty of Medicine, Imperial College London St. Mary’s Hospital London; Division of Medical Sciences and Graduate Entry Medicine, University of Nottingham, Nottingham, UK; NIHR Applied Research Collaboration-East Midlands (ARC-EM), Nottingham, UK; NIHR Nottingham Biomedical Research Centre (BRC), Nottingham, UK; Healthcare Associated Infections Research Group, University of Leeds and Leeds Teaching Hospitals NHS Trust, Leeds, UK; NIHR Leeds In Vitro Diagnostics Co-operative, Leeds Teaching Hospitals NHS Trust, Leeds, UK; NIHR Newcastle In Vitro Diagnostics Co-operative, Translational and Clinical Research Institute, Newcastle University, Newcastle Upon Tyne, UK; Warwick Medical School, University of Warwick, UK; School of Healthcare, University of Leeds, Leeds, UK; NIHR Applied Research Collaboration Yorkshire and Humber, UK; Springfield Healthcare, Leeds, UK; Landermeads Nursing Home, Nottingham, UK; Wren Hall Nursing Home, Selston, UK; Ashmere Nottinghamshire Ltd, Notts, UK; Nuffield Department of Primary Care Health Sciences, University of Oxford, UK; NIHR Community Healthcare MedTech and IVD Co-operative,Oxford,UK

**Keywords:** COVID-19, Point-of-care testing, PCR, care homes

## Abstract

**Introduction:** Reliable rapid testing on COVID-19 is needed in care homes to reduce the risk of outbreaks and enable timely care. Point-of-care testing (POCT) in care homes could provide rapid actionable results. This study aimed to examine the usability and test performance of point of care polymerase chain reaction (PCR) for COVID-19 in care homes.

**Methods:** Point-of-care PCR for detection of SARS-COV2 was evaluated in a purposeful sample of four UK care homes. Test agreement with laboratory real-time PCR and usability and use errors were assessed.

**Results:** Point of care and laboratory polymerase chain reaction (PCR) tests were performed on 278 participants. The point of care and laboratory tests returned uncertain results or errors for 17 and 5 specimens respectively. Agreement analysis was conducted on 256 specimens. 175 were from staff: 162 asymptomatic; 13 symptomatic. 69 were from residents: 59 asymptomatic; 10 symptomatic. Asymptomatic specimens showed 83.3% (95% CI: 35.9%-99.6%) positive agreement and 98.7% negative agreement (95% CI: 96.2%-99.7%), with overall prevalence and bias-adjusted kappa (PABAK) of 0.965 (95% CI: 0.932 – 0.999). Symptomatic specimens showed 100% (95% CI: 2.5%-100%) positive agreement and 100% negative agreement (95% CI: 85.8%-100%), with overall PABAK of 1. No usability-related hazards emerged from this exploratory study.

**Conclusion:** Applications of point-of-care PCR testing in care homes can be considered with appropriate preparatory steps and safeguards. Agreement between POCT and laboratory PCR was good. Further diagnostic accuracy evaluations and in-service evaluation studies should be conducted, if the test is to be implemented more widely, to build greater certainty on this initial exploratory analysis.

**Key points:**

- Point of care tests (POCT) in care homes are feasible and could increase testing capacity for the control of COVID-19 infection.
- The test of agreement between POCT and laboratory PCR for care home residents and the staff was good.
- Adoption of POCT in care homes can be considered with appropriate preparatory steps and safeguards in place.
- Repetitive errors and test malfunctioning can be mitigated with bespoke training for care home staff.
- Integrated care pathways should be investigated to test the high variability of the context of use.

## Introduction

Care home residents have been amongst the worst affected by the COVID-19 pandemic [1]. In England and Wales, the Office of National Statistics estimates that care homes experienced 19,394 excess deaths due to COVID-19 during the first 6 months of 2020 [2].

As the pandemic has continued, attention has turned to how SARS-COV-2 testing might help better protect residents from COVID-19 outbreaks, whilst enabling continuity of service provision and promoting quality of life for residents by restoring routine practices such as family visiting [3].

Point-of-care testing (POCT) describes tests that take place close to the care setting [4, 5]. We have previously described possible roles for POCT for COVID-19 in screening asymptomatic residents and staff, and diagnostic testing for symptomatic residents and staff [6]. For both use cases, POCT could reduce administrative burden and test turnaround times, and reduce the chances of COVID-19 outbreaks through earlier quarantine of positive cases.

The “real world” effectiveness of POCT in care homes is a function of the technical properties of the test (established in laboratory conditions where certainty and parameters are controlled) and its performance in conditions of the uncertainties that exist in homes, such as residents with complex multimorbidity, variation in testing behaviours in staff, variable infection control processes and implementation. POCT should take account of how testing can integrate into existing measures of transmission control, and the complexity involved in preparing for testing, conducting testing, and interpreting and responding to results.

Very little is known as to how well the POCTs can be integrated into care home settings and their readiness for adoption among potential users. If these POCTs are implemented into settings that they have not been designed for there is an inherent risk that the operator will execute the test improperly, use it in the way not intended, or even reject the test for lack of perceived benefits.

As part of the COVID-19 National Diagnostic Research and Evaluation (CONDOR) platform [7], our study aim was to evaluate a point-of-care polymerase chain reaction (PCR) machine for detection of SARS-COV2 in the UK care home setting. Our primary objective was to rapidly explore the usability and readiness of the technology for use in real-world settings, to scope usability requirements, prioritise areas of deployment, and to identify any possible safety concerns and sources of error to be further investigated and mitigated. A secondary objective was to evaluate the agreement between results from the POC test with the laboratory-based Reverse Transcriptase-PCR (RT-PCR), the current benchmark standard for care homes.

## Methods

The technology being evaluated was the POCKIT™ Central Nucleic Acid Analyzer (POCKIT™ Central). This is a benchtop molecular detection system, integrating magnetic bead-based nucleic acid extraction, fluorescence-based insulated isothermal PCR (iiPCR) amplification/detection, and liquid handling technologies to offer a walk-away protocol for nucleic acid detection. The machine can run tests on up to eight specimens at once. Results are displayed on the monitor in less than 1.5 hours. POCKIT™ Central machines were delivered directly to care homes, and training provided, by the manufacturer. Only staff members who received formal training were permitted to use the machine

This study consists of two activities:

1. Scoping usability and use errors.
2. Test agreement with laboratory RT-PCR.

No formal power calculation was undertaken. Further evaluation of diagnostic accuracy is being conducted in other parts of the CONDOR platform.

### Participant recruitment

The CONDOR study has recruited 28 care homes, caring for 1491 residents, to our testing platform. These were recruited through publicity in a national care home WhatsApp COVID-19 peer support group [8], public-facing social media, and the national care home umbrella organisations Care England and the National Care Forum. The care home database is active, and we continue to recruit. It includes nursing and residential homes; corporate chain, independent and third-sector providers; and bed capacity ranging from 20+ -350+ in each home. From this, we selected a purposive sample of four care homes (Table 1), comprising two care homes with nursing and two care homes without nursing (also known as nursing and residential homes respectively), from two regions of the UK, two independents and two small chains, to ensure we maximised our ability to account for differences in staff training, and organisational configuration, that might impact on the implementation or use of a point-of-care test.

**Table 1.** Description of care homes and participants taking part in the study.

| Interviewee's role | Area | Nursing Home | Residential Home | TOT number of beds | Identification Code (IC) |
| --- | --- | --- | --- | --- | --- |
| Care Home Manager | Nottinghamshire | x |  | 70 | CH1 |
| Managing Director | Nottinghamshire |  | X | 140+ | CH2 |
| Project Improvement Officer |  |  |  |  | CH3 |
| Chief Nursing Officer | Oxfordshire | x | X | 350+ | CH4 |
| Managing Director | Nottinghamshire | x |  | 50+ | CH5 |

We chose the number of participants based upon the need to understand workflows around staff and resident routine testing, whilst capturing errors that might occur on repeated use of the machine, by different staff members, on different days, under different circumstances.

### Scoping usability and use errors

To scope usability areas and potential sources of error, semi-structured interviews were conducted with key stakeholders (Table 1). These interviews also enabled further insights regarding potential operational and pathway integration of the new test. Participants were directly involved with trialling POCKIT^™^Central in care homes and had experience in using the device.

Stakeholders were interviewed individually and remotely by a researcher in Human Factors (MM) at the end of the trial. In one instance for reasons of practicality, two interviewees who worked together were interviewed together. Interviews were semi-structured and lasted 30-60 minutes; they were audio and video recorded, with the permission of the participants. Interview schedules focussed on the manufacturer’s instructions for use, how POCKIT^™^Central might be integrated into the diagnostic pathway, the testing strategy and the clinical decision making based on positive and negative results. Participants were then prompted to explore potential usability issues in the use of POCKIT^™^Central (including clarity of test results, potential hazards and the disposal procedure).

### Test agreement with laboratory PCR

We chose to focus primarily on routine staff testing. This is currently conducted weekly, and results from PCR laboratory tests do not always arrive within seven days. A growing body of evidence suggests that asymptomatic transmission from staff is likely to be one of the most important factors in care home outbreaks [9]. We estimated that conducting 65 tests in each of the four care homes would provide adequate information on our objectives. The breakdown of tests between the different groups were: 35 routine staff tests; 15 routine resident tests; and 15 tests for resident or staff who had become symptomatic.

Nasopharyngeal swabs were taken by care home staff, adhering to standard testing procedures recommended by the government [10], using government-issued testing kits. Use of the POCKIT™ Central device followed a Standard Operating Procedure (see Appendix I) written by laboratory staff after risk assessing the extent of viral inactivation in the sample buffer. Once the testing swab had been inserted into the viral transport medium, 200 μL was transferred into the POCKIT™ Central testing well by the care home staff member using a pre-set Gilson pipette and sterile filter tips. The remainder of the specimen was sent for formal laboratory testing as usual. Care home staff documented testing results from the POCKIT™ Central device using a results log, adding formal laboratory results when these returned. Only anonymised data were seen by the research team. Samples were not blinded at any stage.

All test results, including equivocal results and failures, were reported as per US Food and Drug Administration (FDA) guidance. We calculated positive and negative agreement of POCKIT™ Central with the formal laboratory RT-PCR results, the (Cohen) kappa, the Brennan and Prediger statistic (equal to the prevalence and biased adjusted kappa (PABAK)) as well as prevalence based on laboratory RT-PCR results, with their associated 95% confidence intervals. The primary analysis was based on valid results for all tests stratified by symptomatic/asymptomatic participants. These calculations were carried out in Stata/SE 16.1 using: **diagt** and **kappaetc**. We also carried out a sensitivity analysis including all POCKIT™ Central equivocal results and test failures in our calculations described above.

## Results

### Scoping Usability and use errors

The Human Factors experts reviewed all potential critical issues in the main operation tasks of POCKIT^™^Central that could lead to errors and hazards affecting the test operators and the care home residents. Instructions for use and a video demonstration were analysed to highlight the expected source of errors. Four operation tasks and potential risks of use and errors were identified and are outlined in figure 1. Each task and the probability of occurrence of these potential errors were discussed with interview participants.

**Figure 1:**
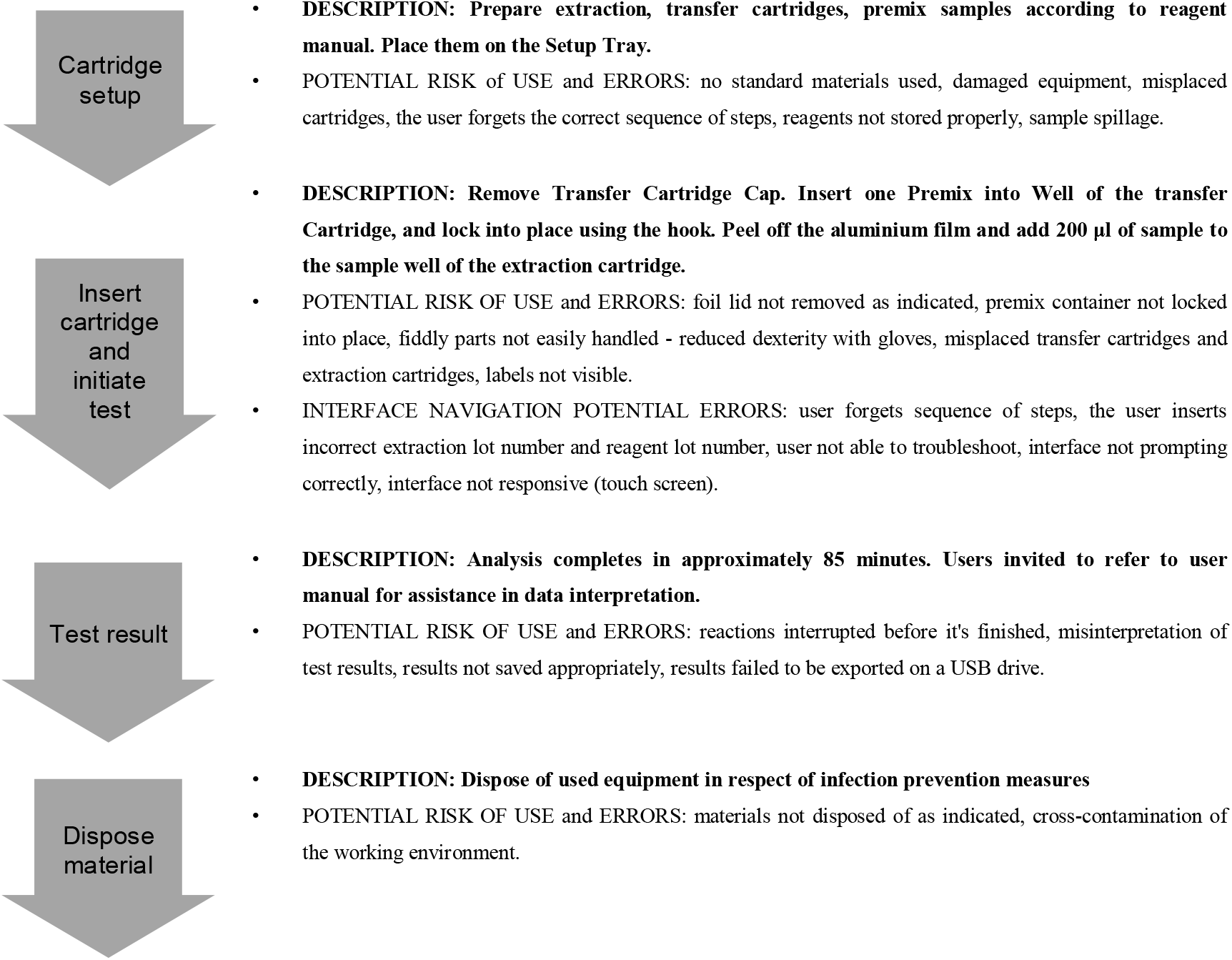
Flowchart of the operative procedure and potential risks-errors of use associated, as highlighted by Human Factors experts

The overall experience with testing equipment was positive. Using the device did not cause excessive cognitive workload as users easily remembered the required steps. Handling components and inserting them into the correct location was not problematic and did not lead to errors. The test activation – which included navigating the digital interface – was considered “easy to manage” (CH2-CH3) and staff members appreciated the presence of prompts and a home button for better control of the process (CH1). No results misinterpretation was observed. Results were correctly recorded. Each test was linked to a unique ID and the data automatically saved for retrieval.

The list of observed errors and unexpected situations have been divided based on the four operative procedure’s stages. Relevant quotes from participants and mitigation strategies to reduce the occurrence of errors are reported (Table 2). Quotes are selected for their relevance to inform key usability aspects and to corroborate findings.

**Table 2.**
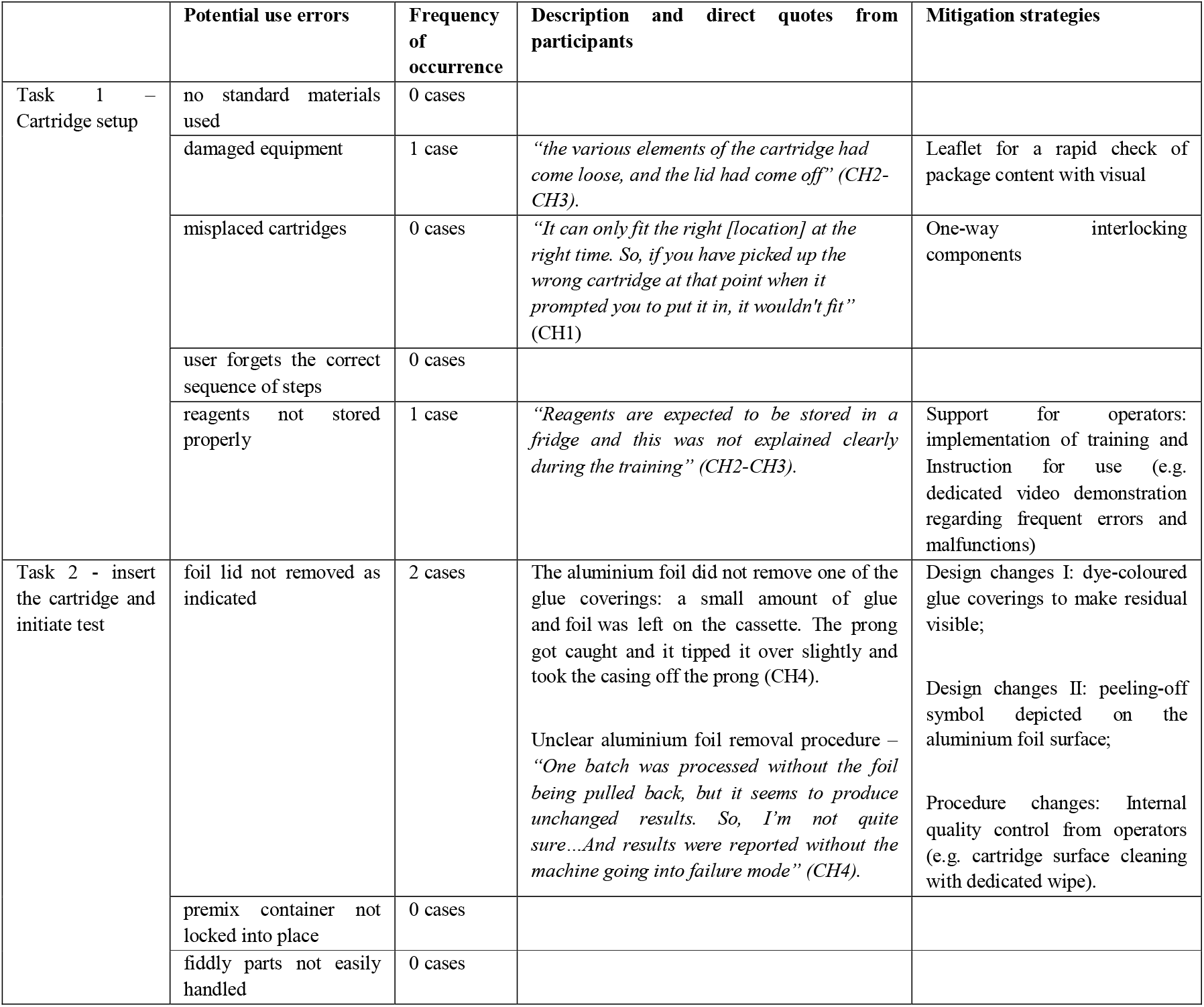
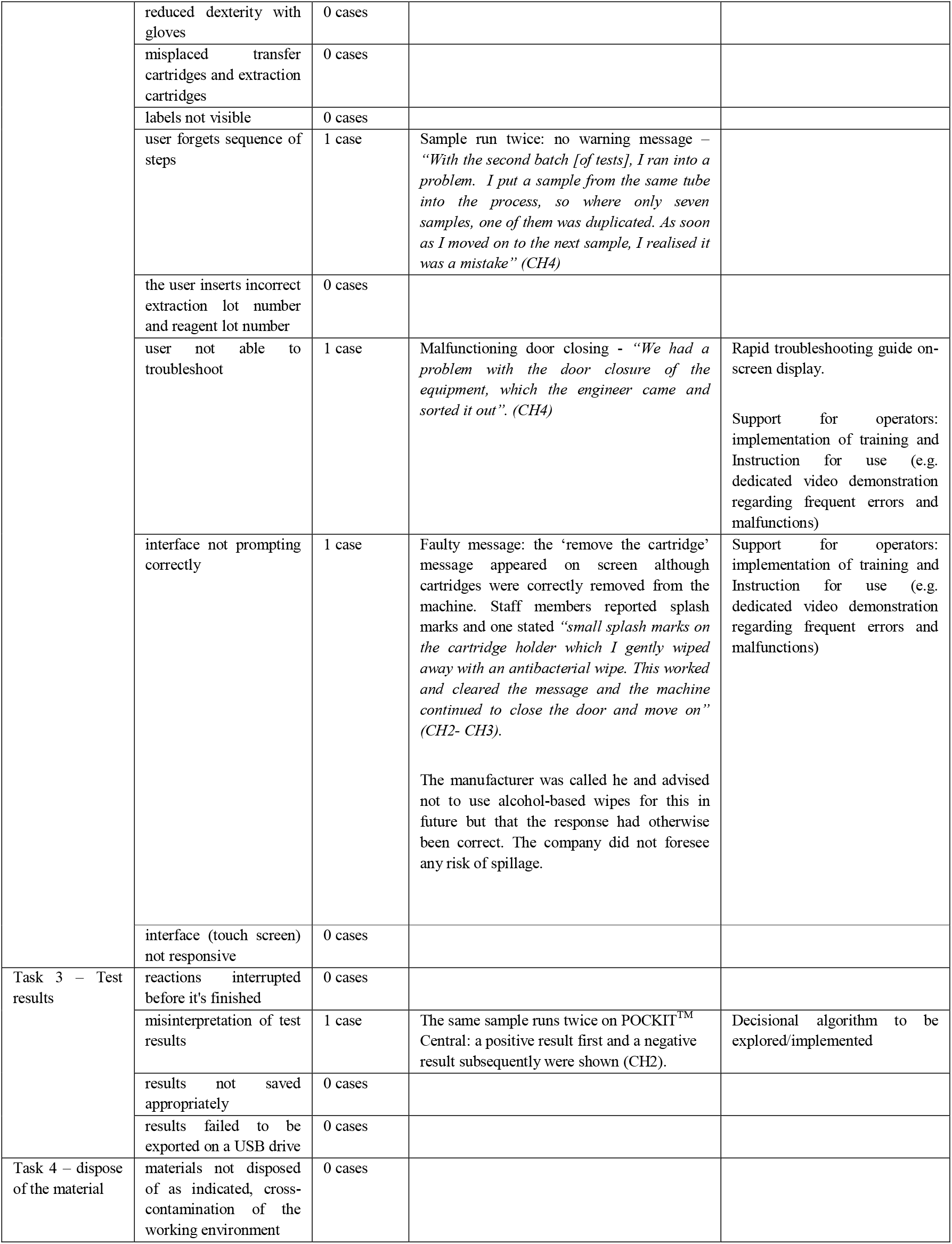
List of observed errors of use and mitigation strategies.

|  | Potential use errors | Frequency of occurrence | Description and direct quotes from participants | Mitigation strategies |
| --- | --- | --- | --- | --- |
| Task 1 – Cartridge setup | no standard materials used | 0 cases |  |  |
|  | damaged equipment | 1 case | *“the various elements of the cartridge had come loose, and the lid had come off” (CH2-CH3).* | Leaflet for a rapid check of package content with visual |
|  | misplaced cartridges | 0 cases | *“It can only fit the right [location] at the right time. So, if you have picked up the wrong cartridge at that point when it prompted you to put it in, it wouldn't fit” (CH1)* | One-way interlocking components |
|  | user forgets the correct sequence of steps | 0 cases |  |  |
|  | reagents not stored properly | 1 case | *“Reagents are expected to be stored in a fridge and this was not explained clearly during the training” (CH2-CH3).* | Support for operators: implementation of training and Instruction for use (e.g. dedicated video demonstration regarding frequent errors and malfunctions) |
| Task 2 - insert the cartridge and initiate test | foil lid not removed as indicated | 2 cases | The aluminium foil did not remove one of the glue coverings: a small amount of glue and foil was left on the cassette. The prong got caught and it tipped it over slightly and took the casing off the prong (CH4).<br>Unclear aluminium foil removal procedure – *“One batch was processed without the foil being pulled back, but it seems to produce unchanged results. So, I'm not quite sure...And results were reported without the machine going into failure mode” (CH4).* | Design changes I: dye-coloured glue coverings to make residual visible;<br>Design changes II: peeling-off symbol depicted on the aluminium foil surface;<br>Procedure changes: Internal quality control from operators (e.g. cartridge surface cleaning with dedicated wipe). |
|  | premix container not locked into place | 0 cases |  |  |
|  | fiddly parts not easily handled | 0 cases |  |  |
|  | reduced dexterity with gloves | 0 cases |  |  |
|  | misplaced transfer cartridges and extraction cartridges | 0 cases |  |  |
|  | labels not visible | 0 cases |  |  |
|  | user forgets sequence of steps | 1 case | Sample run twice: no warning message – *“With the second batch [of tests], I ran into a problem. I put a sample from the same tube into the process, so where only seven samples, one of them was duplicated. As soon as I moved on to the next sample, I realised it was a mistake” (CH4)* |  |
|  | the user inserts incorrect extraction lot number and reagent lot number | 0 cases |  |  |
|  | user not able to troubleshoot | 1 case | Malfunctioning door closing - *“We had a problem with the door closure of the equipment, which the engineer came and sorted it out”. (CH4)* | Rapid troubleshooting guide on-screen display.<br>Support for operators: implementation of training and Instruction for use (e.g. dedicated video demonstration regarding frequent errors and malfunctions) |
|  | interface not prompting correctly | 1 case | Faulty message: the ‘remove the cartridge’ message appeared on screen although cartridges were correctly removed from the machine. Staff members reported splash marks and one stated *“small splash marks on the cartridge holder which I gently wiped away with an antibacterial wipe. This worked and cleared the message and the machine continued to close the door and move on” (CH2- CH3).*<br>The manufacturer was called he and advised not to use alcohol-based wipes for this in future but that the response had otherwise been correct. The company did not foresee any risk of spillage. | Support for operators: implementation of training and Instruction for use (e.g. dedicated video demonstration regarding frequent errors and malfunctions) |
|  | interface (touch screen) not responsive | 0 cases |  |  |
| Task 3 – Test results | reactions interrupted before it's finished | 0 cases |  |  |
|  | misinterpretation of test results | 1 case | The same sample runs twice on POCKIT™ Central: a positive result first and a negative result subsequently were shown (CH2). | Decisional algorithm to be explored/implemented |
|  | results not saved appropriately | 0 cases |  |  |
|  | results failed to be exported on a USB drive | 0 cases |  |  |
| Task 4 – dispose of the material | materials not disposed of as indicated, cross-contamination of the working environment | 0 cases |  |  |

### Integration pathway

Further comments covered the integration aspect of POCT in a care home setting. Overall, participants reacted enthusiastically to the possibility of its implementation as part of their care home’s testing capability.

To run tests correctly, to safeguard operators and residents, and to protect the testing equipment from damage, a dedicated room must be set up with operators wearing fresh PPE. As CH4 explained: *“I do think we need a dedicated room, which remains a dedicated room for long. It is because it’s a heavy machine. And you would not be transporting it from place to place (risk of damages) […] Dedicated room (that serves all the different houses) made into the testing room, separate from homes, outside the community (no access to residents)”*.

Two ways in which the test results might inform practice were discussed: i) the diagnostic test for symptomatic residents and staff; ii) the screening test for asymptomatic residents and staff.

As a one-off test, POCKIT™ Central was considered by staff to be appropriate for testing symptomatic residents and for obtaining a rapid result to enable timely care decisions: *“I can see a huge advantage because for those people that are symptomatic suddenly; I think it could solve a lot of situations for which you see an immediate answer, or a more immediate answer would be useful to complement it on it”* (CH1).

A rapid test would better inform isolation decisions, avoiding unnecessary segregation, with benefits to residents’ wellbeing and the wider care home community: *“when we’ve had people who were symptomatic, rather than having to close the community we would now put in all the precautions without stopping relatives coming in […] we only have to have two suspected or two confirmed cases”* (CH4). This technology would contribute to maintaining a testing momentum in care home facilities, and it would provide a consistent diagnostic approach. Having care home managers in control would relieve the burden of managing unpredictable factors, including frequent changes of government strategy around testing and isolation: *“And this would allow us to test people and keep the workforce in a constant continuity for the people in our care. It offers reassurance to the staff”* (CH4).

However, the main concern of staff was the test turnaround of 85 minutes, as they considered it would require extra staff capacity to test the whole home in approximately two days. Given the test turnaround time, we identified considerable staffing issues with asymptomatic cases as these will require significant amounts of staff time: *“If you’re wanting to replace testing here with something like that, the capacity of it in the time it would take to get through the number of people we need to get to. Which would be a huge problem here”* (CH1).

In big care homes (200+), it might take more than two days to screen all the residents if Point-of-Care tests were deployed in this context. The staff in all care homes outnumbered the residents, and are currently subject to weekly testing, implying even greater workload. Consideration should be given to the resource implications for care homes. There was some disagreement amongst respondents about whether this would essentially mirror the staffing requirements for existing routine screening tests, or whether it would demand extra staff time.

### Agreement with laboratory RT-PCR

In total 278 tests were run. This was slightly more than originally requested as care homes had more test kits delivered from the manufacturer due to how reagents were batched. Tests were carried out on 252 asymptomatic participants (176 staff and 76 residents) and 26 symptomatic participants (13 staff and 13 residents) (Table 3 and Appendix II Table A1 stratified by staff/residents). Formal laboratory results were indeterminate for five specimens. POCKIT^™^Central results were not available for 17 tests; one was a sporadic error, the remaining 16 were compromised by sealing glue left on top of the POCKIT^™^Central cartridge. This was an avoidable error, which is described in detail above. A Standard for Reporting of Diagnostic Accuracy Studies (STARD) Flow diagram is shown in Figure 2.

**Table 3.** Full results from POCKIT and Laboratory RT-PCR.

Asymptomatic cases
|  | Laboratory (PCR) |  |  |  |
| --- | --- | --- | --- | --- |
| POCKIT | Negative | Positive | Uncertain/error | Total |
| Negative | 222 | 1 | 4 | 227 |
| Positive | 3 | 5 | 0 | 8 |
| Uncertain/error | 17 | 0 | 0 | 17 |
| Total | 242 | 6 | 4 | 252 |

Symptomatic cases
|  | Laboratory (PCR) |  |  |  |
| --- | --- | --- | --- | --- |
| POCKIT | Negative | Positive | Uncertain/error | Total |
| Negative | 24 | 0 | 1 | 25 |
| Positive | 0 | 1 | 0 | 1 |
| Uncertain/error | 0 | 0 | 0 | 0 |
| Total | 24 | 1 | 1 | 26 |

**Figure 2.**
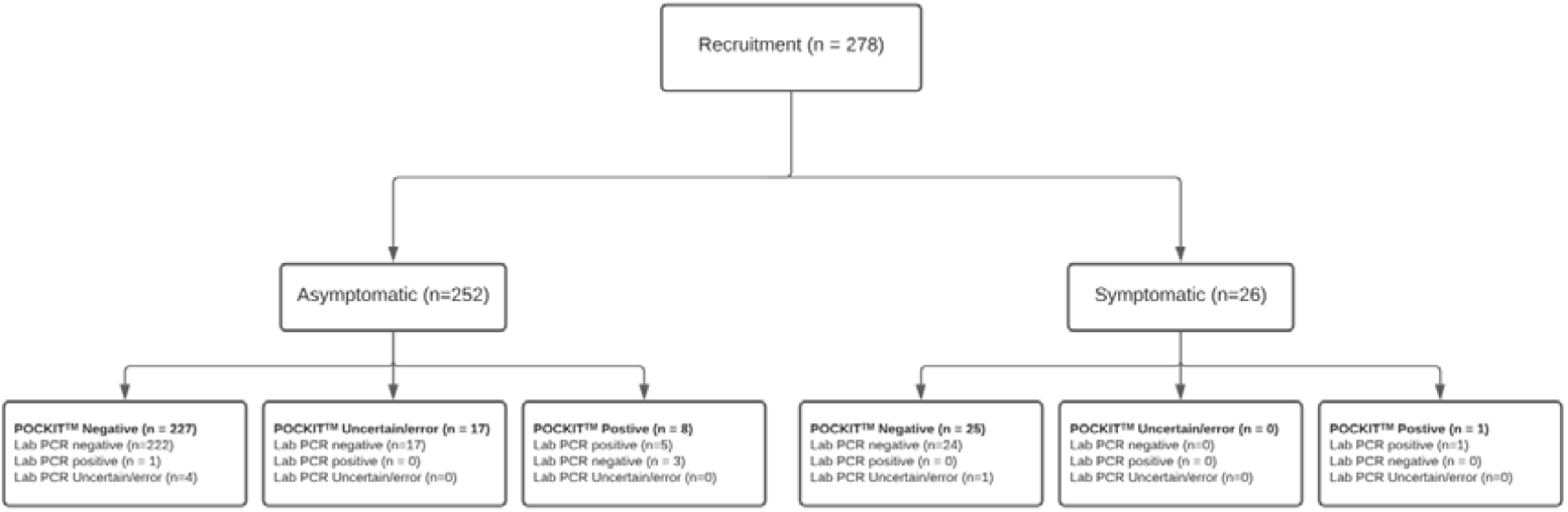
Standards for Reporting of Diagnostic Accuracy Studies (STARD) Flow diagram.

Thus, agreement analysis was conducted on 256 specimens: 177 from staff, 164 asymptomatic, and 13 symptomatic; 79 from residents, 67 asymptomatic and 12 symptomatic. Estimated prevalence data, positive and negative agreement, and kappa and PABAK results for asymptomatic and symptomatic participants are shown in Table 4. Equivalent statistics based on including all POCKIT™ Central equivocal and failures are reported in Appendix II Table A2.

**Table 4.** Agreement based on valid measures.

Asymptomatic cases
|  | LAB -ve | LAB +ve | Total |
| --- | --- | --- | --- |
| POCKIT negative | 222 | 1 | 223 |
| POCKIT positive | 3 | 5 | 8 |
| Total | 225 | 6 | 231 |
| Test attributes |  | (95% CIs) |  |
| Prevalence (LAB measure) | 2.6% | (0.96%-5.57%) |  |
| Positive agreement (LAB positive as denom) | 83.3% | (35.9%-99.6%) |  |
| Negative agreement (LAB negative as denom) | 98.7% | (96.2%-99.7%) |  |
| Kappa | 0.706 | (0.429-0.982) |  |
| Prevalence and bias-adjusted Kappa (PABAK) | 0.965 | (0.932-0.999) |  |

Symptomatic cases
|  | LAB -ve | LAB +ve | Total |
| --- | --- | --- | --- |
| POCKIT negative | 24 | 0 | 24 |
| POCKIT positive | 0 | 1 | 1 |
| Total | 24 | 1 | 25 |
| Test attributes |  | (95% CIs) |  |
| Prevalence (LAB measure) | 4% | (1%-20.4%) |  |
| Positive agreement (LAB positive as denom) | 100% | (2.5%-100%) |  |
| Negative agreement (LAB negative as denom) | 100% | (85.8%-100%) |  |
| Kappa | 1 | (1-1) |  |
| PABAK | 1 | (1-1) |  |

## Discussion

This paper reports the first exploratory in-context evaluation of point-of-care (POC) PCR for SARS-COV-2 in the care home setting. We found no significant hazards and the sources of error identified were amenable to relatively minor changes in training or test workflow. Therefore, adopting POCKIT^™^Central in a care home setting can be considered safe and feasible with appropriate preparatory steps and safeguards in place.

The positive and negative agreement reported in this study can be compared to the measures of sensitivity and specificity based on RT-PCR as the reference standard. The estimates presented here for POCKIT™ Central were acceptable, particularly for symptomatic participants. Care home staff in our sample became very enthusiastic that the test might also be deployed to screen care home visitors for COVID-19, as part of the effort to re-open care homes to visitors. However, the specific workflow or safety issues associated with testing visitors were not investigated in this study.

The technology, as expected for PCR, shows much higher accuracy, based upon the level of positive agreement seen, than the lateral flow devices recently deployed as part of the UK mass testing programme [11]. PCR is likely to be substantially more expensive in terms of up-front costs than using lateral flow tests. It does, though, potentially represent a single step testing solution, shifting expense from laboratories to the community. This would offer significant resource savings for care homes in comparison with the multi-stage testing approaches which might be needed if using currently available lateral flow tests. The wider work of the CONDOR programme involves care pathway mapping which is informing NICE (National Institute for Health and Care Excellence) exploratory economic modelling of SARS-CoV-2 viral detection POCTs [12]. This will consider the cost of novel POCTs as compared with standard laboratory testing alongside the potential benefits accumulated.

An important practical consideration before any roll-out of this technology, is that the CE mark for the POCKIT™ Central, by which the Medicines and Healthcare Products Regulatory Agency (MHRA) defines its terms of use, restricts its use to “healthcare professionals”. Although some are registered nurses, many care home staff are not registered healthcare professionals. The CE mark would have to be modified to take this into account. This is not likely to be confined to POCKIT™ Central and is a reflection of the fact that POC tests have not hitherto been widely deployed in care homes. If such technology is to be widely deployed, then CE marks will have to take account of the competency mix of care home staff.

A limitation of this study, which it is important to recognise, is that it was not statistically powered as a diagnostic accuracy study. The low number of positives seen explains the wide confidence intervals reported for all estimates, particularly positive agreement. We report several measures of agreement stratified by symptomatic/asymptomatic to reflect potential different use of these tests. These statistics have been shown to have limitations and biases but can serve as an indication of overall agreement between these tests, hence our reporting of multiple measures alongside full test results. Our sensitivity analysis to explore the impact of POCKIT™ Central uncertain results and machine errors showed our results to be robust.

Another limitation is the relatively small sample of care homes involved. Although we sought to evaluate the technology in a purposive sample of homes that represented the range of training and registration across UK care homes more generally, the organisational configuration of UK care is recognised to be highly heterogeneous. Also, the “early adopter” care homes that routinely engage with research or service development by academics might be atypical. If wider roll-out of POCKIT™ Central in care homes were to be considered as a consequence of this evaluation, we would recommend doing so in a staged way, with further in-service evaluation studies to tailor training/training material for care homes staff and to fully assess how POCKIT™ Central can be integrated into the full range of care home workflows. A robust statistically powered diagnostic accuracy study conducted in parallel could provide conclusive evidence as to whether POCKIT™ Central compared to laboratory RT-PCR performs as well in the care home setting as it does in other contexts.

In conclusion, this evaluation of POCKIT™ Central highlights that POCT PCR for COVID-19 settings is feasible in care home settings. We have not, hitherto, identified significant safety concerns. The agreement with diagnostic benchmark laboratory PCR tests was good. If implemented at greater scale, further diagnostic accuracy evaluations alongside a further evaluation of usability and integration should be conducted in parallel. This could include evaluation of additional use cases, including extending testing to care home visitors.

## Data Availability

The authors confirm that the data supporting the findings of this study are available within the article and its supplementary materials.
The data that support the usability study are available on request from the corresponding author, [AG]. The data are not publicly available due to restrictions regarding the privacy of research participants.

## Ethical approval

This project was approved as a Service Evaluation by Imperial College Healthcare NHS Trust (ICHNT) – registration no. 471.

## Acknowledgements

The authors would like to thank care home managers and staff members who took part in the study and members of the CONDOR platform for their comments: Mr Graham Prestwich and Ms Val Tate.

## Appendix I

Standardised Operating Procedure for Use of HORIBA POCKIT™Central ™ for Testing of COVID-19

This guide will provide you with step by step instructions on how to successfully use the Horiba POCKIT™ to test individuals for COVID-19. Please ensure the steps are followed to enable the test to be completed in the correct manner for the best chance of an accurate result.

### Health and Safety

- When handling samples, full PPE must be worn at all times for your protection and the protection of others. This includes gloves, gown, face masks and visors (if applicable).
- Once a swab is taken from a resident, immerse the swab in the viral transport media in the bottom of the tube, and push down so that the cap at the end of the swab is securely in place.
- Samples can be run in batches of 8 samples therefore do not need to be run straight away if more than 1 sample is set to be collected.
- For any batch collecting, please keep samples to one side on paper towels in the testing room away from any waste bins or areas that may be contaminated.
- Tests must be processed in a separate, designated room with all PPE fully worn and consumables disposed in waste streams provided.
- When testing is performed, only one person must be allowed into the room at any time
- Testing room should be secured at all times with the door kept closed
- Test preparation should be conducted away from the waste bins but still in the same room. The liquid solutions will contain amplified RNA therefore there is a potential risk of contamination to other samples and work surfaces
- Take care not to spill any of the liquid from the tubes. If a spillage does occur, Clean up any spillage with any disinfectant wipes such as Clinell wipes or any other 0.1% hypochlorite solution
- Should any spillage occur from the extraction cartridge, this must be cleaned with Isopropanolol such as a 70% Isopropyl alcohol solution (Extraction buffer contains guanidine therefore SHOULD NOT be cleaned with hypochlorite)
- Ensure all surfaces are cleaned down thoroughly with a detergent disinfectant suitable for removing SARS-CoV-2 this would also include 0.1% hypochlorite solution or clinell wipes

### Training

Only members of staff that are trained to use the Horiba POCKIT™ will be able to carry out testing.

First Use only:

1. Upon arrival of instrument, plug into mains supply, ensure the air filter at the back of the analyser is not blocked in anyway.
2. Ensure the door at front of analyser will not be blocked from opening.
3. Once analyser is plugged in and switched on, the home screen will prompt for removal of ‘Transfer Cartridges’.
4. The screen will state ‘Door Opening’ - Press ‘OK’ and the door will open to expose the cartridge tray.
5. Remove the cartridges in slot A+H, close the door and press ‘End’ to take you to the home screen.
6. Instructions for setting up the machine can be found in the manufacturer instructions

### Swab collection

1. Wearing full PPE, obtain nasal and throat swab from each patient that requires a COVID test
2. Swab the patient at the bedside (or their seat) and then place the swab into the tube provided with swab
3. This tube will contain a liquid solution at the bottom, ensure the swab sits fully into the liquid at the bottom of the swab transport tube before closing securely. (The large tubular end of the swab is the lid)
4. Swirl the swab and liquid solution to ensure any materials from the swab is immersed fully into the liquid.
5. You will see the liquid turn slightly cloudy
6. Take swabs to the secure testing room
7. Before entering the room, all PPE must be discarded into separate clinical waste bins with a lid.
8. Put on fresh PPE upon entry into the testing room. DO NOT enter the testing room with the same PPE

### Running a Test

1. When entering the testing room, ensure the door is closed behind you
2. Place the swabs on a paper towel on a clean work surface next to the analyser
3. Gather all materials required for the test.
  a. Pipette (set at 200ul)
  b. Sterile, filtered, Pipette tips (keep them in the box)
  c. Extraction Cartridge (one per test)
  d. Reagent (transfer) cartridge (one per test)
  e. Primer mix vial if included separately (one per test)
4. On the Home screen on the analyser, press ‘Start’ to begin a new test
5. You will then be asked to type in the extraction cartridge lot number, reagent lot number (transfer cartridge) and a sample ID.
6. To enter sample ID click on the relevant box and type in the information
7. Select the Extraction Lot No and type this, this can be found on the cartridge packaging - repeat for reagent ID
8. The door will be open
9. Remove the extraction cartridge from its aluminium packaging, with the notched side face towards you, place into slot A on the setup tray (left hand side, the slot closest to you)
10. Remove the film
11. Using the pipette provided, take a filter pipette tip from the tip box by pressing the pipette down onto the opening of the tip to ensure its firmly in place
12. Open the tube containing the swab, place the pipette into the tube with the tip touching the liquid
13. Press down the button on the pipette and lift the button slowly, you will see liquid filling into the pipette tip. The pipette will stop filling once the button is back into its raised position
14. Take the pipette out of the tube and place it at the opening of the first well on the extraction cartridge, press the button fully down to ensure all the liquid leaves the pipette tip and goes into the well
15. Dispose of the pipette tip into a sharps bin by pressing the eject button on the pipette directly into the opening of the sharps bin
16. Next, type in the reagent lot number (transfer cartridge) which can be found on the foil packaging
17. Open the lid of the transfer cartridge and slot the cartridge into the back right hand column position A (next to extraction cartridge) on the set-up tray. The notch part of the cartridge should be facing away from you when you slot it in
18. Place a tube of the premix vial into the third well on the cartridge. The plastic seal on the vial must not be removed
19. Once the reagent cartridge is in place, the system will match up the lot number and cartridge automatically (you don’t need to do anything). If this is the only test you are doing press ‘Confirm’. This will bring up the ‘Run Overview’ page
20. If you have more than one sample to test, on the screen press ‘Next’ to repeat steps 10-19 again for the next patient sample. Repeat these steps for as many samples (up to 8 and columns B-H) that require testing at that time
21. Once all samples are loaded, press ‘Confirm’, this will take you to the ‘Run Overview’ Screen.
22. Once you have checked all information is correct, press ‘Run’.
23. The screen will prompt that the door is closing, press ‘OK’
24. The test will run for 85 minutes

### To View Results

The timer remains on the screen and counts down until the testing is completed, the results are then displayed in the same page.

Once the test is completed, follow the onscreen steps to remove the cartridges. The extraction cartridge must be placed into a lidded clinical waste bin (kept closed when not in use) while the reaction cartridge (transfer cartridge) must be disposed of in a sharps bin.

The interior of the analyser should be cleaned with warm water and paper towels ONLY.

DO NOT use alcohol wipes to clean the interior of the analyser. Keep all alcohol wipes away from the sample tray when the door is open

Following the completion of testing, all surfaces must be wiped down with appropriate disinfectant (clinell wipes or hypochlorite solution).

Full waste bins must be sealed, disposed of, waste bags replaced, and the room kept secure upon exit.

Please wash your hands thoroughly with soap and water after PPE is removed

## Appendix II

**Table A1.**
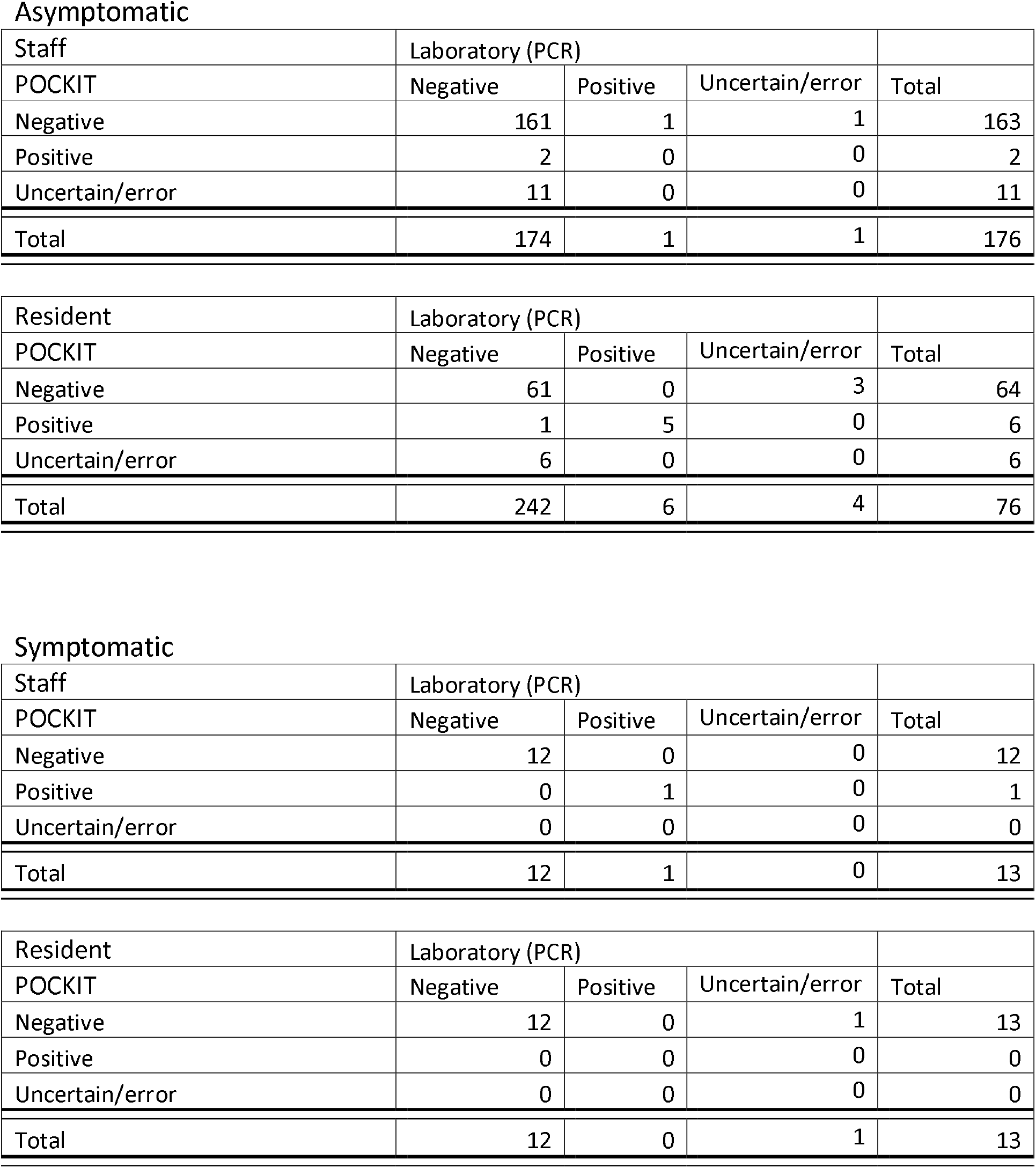
Full results from POCKIT and Laboratory RT-PCR stratified by Staff vs. Residents.

**Table A2.** Agreement based on valid measures for laboratory including all POCKIT results.

Asymptomatic
|  | LAB -ve | LAB +ve | Total |
| --- | --- | --- | --- |
| POCKIT -ve | 222 | 1 | 223 |
| POCKIT +ve | 3 | 5 | 8 |
| POCKIT uncertain/machine error | 17 | 0 | 17 |
| **Total** | **242** | **6** | **248** |
| Test attributes |  | (95% CIs) |  |
| Prevalence (LAB measure) | 2.4% | (0.89%-5.19%) |  |
| Positive agreement (LAB+ as denom) | 83.3% | (35.9%-99.6%) |  |
| Negative agreement (LAB- as denom) | 91.7% | (87.5%-94.9%) |  |
| Kappa | 0.305 | (0.095-0.514) |  |
| PABAK | 0.873 | (0.821-0.925) |  |

Symptomatic
|  | LAB -ve | LAB +ve | Total |
| --- | --- | --- | --- |
| POCKIT -ve | 24 | 0 | 24 |
| POCKIT +ve | 0 | 1 | 1 |
| POCKIT uncertain/machine error | 0 | 0 | 0 |
| **Total** | **24** | **1** | **25** |
| Test attributes |  | (95% CIs) |  |
| Prevalence (LAB measure) | 4% | (1%-20.4%) |  |
| Positive agreement (LAB+ as denom) | 100% | (2.5%-100%) |  |
| Negative agreement (LAB- as denom) | 100% | (85.8%-100%) |  |
| Kappa | 1 | (1-1) |  |
| Prevalence adjusted Kappa | 1 | (1-1) |  |

## References

1. Devi R, Hinsliff-Smith K, Goodman C, et al.; The COVID-19 Pandemic in UK Care Homes-Revealing the Cracks in the System. J Nurs Home Res 2020;6:58–60.

2. Office of National Statistics. Deaths involving COVID-19 in the care sector, England and Wales: deaths occurring up to 12 June 2020 and registered up to 20 June 2020 (provisional). https://www.ons.gov.uk/peoplepopulationandcommunity/birthsdeathsandmarriages/deaths/a. (28 October 2020, date last accessed)

3. British Geriatrics Society. COVID-19: Managing the COVID-19 pandemic in care homes for older people. 2020. (12/10/2020, date last accessed)

4. Gutierres SL, Welty TE; Point-of-care testing: an introduction. Annals of Pharmacotherapy 2004;38(1):119–125.

5. Nguyen T, Chidambara VA, Andreasen SZ, et al.; Point-of-Care Devices for Pathogen Detections: The Three Most Important Factors to Realize towards Commercialization. TrAC Trends in Analytical Chemistry 2020:116004.

6. Micocci M, Gordon A, Allen J, et al.; Understanding COVID-19 testing pathways in English care homes to identify the role of point-of-care testing: an interview-based process mapping study. medRxiv 2020.

7. COVID-19 National Diagnostic Research and Evaluation (CONDOR) platform. https://www.condor-platform.org/ (23/11/2020, date last accessed)

8. Spilsbury K, Devi R, Griffiths A, et al.; SEeking AnsweRs for Care Homes during the COVID-19 pandemic (COVID SEARCH). Age and Ageing 2020.

9. UK Department of Health and Social Care. Vivaldi 1: Coronavirus (COVID-19) care homes study report. https://www.gov.uk/government/statistics/vivaldi-1-coronavirus-covid-19-care-homes-study-report (12/10/2020, date last accessed)

10. Public Health England. COVID-19: Guidance for Taking Swab Samples. https://www.gov.uk/government/publications/covid-19-guidance-for-taking-swab-samples (14/11/2020, date last accessed)

11. Preliminary report from the Joint PHE Porton Down & University of Oxford SARS-CoV-2 test development and validation cell: Rapid evaluation of Lateral Flow Viral Antigen detection devices (LFDs) for mass community testing. https://www.ox.ac.uk/sites/files/oxford/media_wysiwyg/UK%20evaluation_PHE%20Porton%20Down%20%20University%20of%20Oxford_final.pdf date last accessed)

12. National Institute for Health and Care Excellence. Exploratory economic modelling of SARS-CoV-2 viral detection point of care tests and serology tests. https://www.nice.org.uk/guidance/indevelopment/gid-dg10038/documents (23/11/2020, date last accessed)

